# The remaining unknowns: A mixed methods study of the current and global health research priorities for COVID-19

**DOI:** 10.1101/2020.06.24.20138198

**Authors:** Alice Norton, Arancha De La Horra Gozalo, Nicole Feune de Colombi, Moses Alobo, Juliette Mutheu Asego, Zainab Al-Rawni, Emilia Antonio, James Parker, Wayne Mwangi, Colette Adhiambo, Kevin Marsh, Marta Tufet Bayona, Peter Piot, Trudie Lang

## Abstract

**Introduction:** In March 2020 the World Health Organisation (WHO) released a Global Research Roadmap in an effort to coordinate and accelerate the global research response to combat COVID-19 based on deliberations of 400 experts across the world. Three months on, the disease and our understanding have both evolved significantly. As we now tackle a pandemic in very different contexts and with increased knowledge, we sought to build on the work of the WHO to gain a more current and global perspective on these initial priorities.

**Methods:** We undertook a mixed methods study seeking the views of the global research community to i) assess which of the early WHO roadmap priorities are still most pressing; ii) understand whether they are still valid in different settings, regions or countries; and iii) identify any new emerging priorities.

**Results:** Thematic analysis of the significant body of combined data shows the WHO roadmap is globally relevant, however, new important priorities have emerged, in particular, pertinent to low and lower-middle income countries (less resourced countries), where health systems are under significant competing pressures. We also found a shift from prioritising vaccine and therapeutic development towards a focus on assessing the effectiveness, risks, benefits and trust in the variety of public health interventions and measures. Our findings also provide insight into temporal nature of these research priorities, highlighting the urgency of research that can only be undertaken within the period of virus transmission, as well as other important research questions but which can be answered outside the transmission period. Both types of studies are key to help combat this pandemic but also importantly to ensure we are better prepared for the future.

**Conclusion:** We hope these findings will help guide decision making across the broad research system including the multi-lateral partners, research funders, public health practitioners, clinicians and civil society.

**Summary box:** *What is already known?:* The WHO produced a roadmap that set out the research priorities following a meeting in February, just before COVID-19 was declared a Pandemic. Now, at this point in the evolution of this novel disease across the world, and almost 6 months later, it is important to assess whether these priorities remain and if research teams in all countries across the globe agree that these are the most important question that need to be tackled within their health care setting and communities, both to mitigate this outbreak and to learn for next time.

*What are the new findings?:* Over 3,000 healthcare workers and researchers contributed to this research and their data tells us that across the globe there has been a shift in priorities and new questions have emerged, particularly from low-resourced settings. For example, there is a strong call for evidence on the relative effectiveness and optimal implementation of public health interventions in varied global settings, for social science studies to guide how to gain public trust and mitigate myths, to understand the impact on already present diseases within communities, and to explore the ethics of research within a pandemic.

*What do the new findings imply?:* The WHO roadmap is globally relevant, however, our findings also provide insight into the temporal nature of these research priorities, highlighting the urgency of research that can only be undertaken within the period of virus transmission, as well as other important research questions but which can be answered outside the transmission period. Both types of studies are key to help combat this pandemic but also importantly to ensure we are better prepared for the future.

## Introduction

COVID-19 was declared a public health emergency of international concern on 30^th^ January 2020 (1) and then a global pandemic on 11^th^ March 2020.(2) The World Health organisation (WHO) published their Global Research Roadmap (3) on 12^th^ March 2020, within the context of the situation and the epicentre of infection at that time. The Roadmap was built on deliberations of the Global Research Forum, whereby over 400 participants from different sectors across the world, identified 3-4 immediate research priorities for the following three months across each of 9 themes.

Now, in June 2020 we see the evolution of this pandemic at different points across the globe. We know from our previous experience with Ebola and other outbreaks (4, 5) that it is essential to embed research into the response to an outbreak, and that there is a finite and unknown window where these questions can be answered. COVID-19 is an unprecedented situation and therefore we must take every opportunity to undertake all the possible research that funding and capabilities allow; and high-quality studies should happen everywhere there are cases in order to maximise the evidence generated and ensure that the resultant data and findings are globally applicable. Therefore, it is important to assess now, what are the most key remaining global health questions that need to be addressed, both to ensure this pandemic can be halted and to learn for future outbreaks of this pathogen or another.

This research intentionally builds from the WHO Roadmap, with the aim of strengthening the global health research response effort already aligned to this, rather than generating a completely new set of priorities. Using broad consultative workshops, we have identified additional considerations beyond the WHO Roadmap scope, in order to broaden the current global research priorities at this point in time to tackle the COVID-19 pandemic and to help learn for any future outbreaks.

## Methods

An online multi-language survey was developed where ranking questions were coupled with open-ended questions. This was based on a previous survey led by the African Academy of Science (AAS) that was undertaken in March 2020 to assess how well the WHO priorities were applicable to Africa.(6) Here we worked from the AAS survey so we could now assess whether the findings remained relevant across the globe, and if they had changed over time. Seventy-three potential priorities (41 from the original WHO document and 32 generated as part the AAS survey and consultations) were arranged under the nine topic headings used in the WHO Research Roadmap. Participants ranked their top three options for both short- and long-term priorities (18 total ranking questions). Free text boxes were provided under each of the broad topics, where participants were asked to list any research priority they felt was not included in the options provided. Recognising that this survey inherently focussed respondents on the existing WHO priority framework, we expanded our consultation through workshops to enable broader discussions of research priorities.

After the survey closed a virtual workshop was held on the 5^th^ of June to seek wider global comment and discussion on the survey findings and to discuss current priorities and unmet research areas beyond the scope of the existing WHO priority framework. We conducted ten further open access workshops with research teams and health workers across the globe, led by the The Global Health Network (TGHN) COVID-19 Research Implementation and Knowledge Hub between 14th April and 12th June 2020. These workshops meetings were recorded with permission of participants, and comments and questions captured. A thematic content analysis methodology was developed to report the findings of each.(7) Here we applied this to the cumulative data of all 11 workshops to add to the survey data and better address the question: *what are the current global research priorities during the COVID19 pandemic?*

### Quantitative Data Analysis Methods

Responses from the survey were download in excel format, all data was fully anonymised, password protected and access restricted to the study team. Descriptive analyses were undertaken within excel to provide a ranking score for each research priority for immediate and longer term, as per the survey. Priorities ranked as first were given a score of 3, those ranked second were given a score of 2 and those ranked third were given a score of 1. This analysis was conducted within the category headings from the WHO roadmap and included both the original WHO priorities and new priorities suggested in the AAS report. Therefore, these data show us how responders currently rank the priorities set within the WHO roadmap and the AAS report. The data were split for comparison between the global researcher responses and those originating from less-resourced settings. Within the less-resourced setting category, we include low and lower-middle countries as defined by the World Bank.

### Qualitative Data Analysis Method

The aim of the open-ended survey was to determine whether there are new priorities that were not included in the original WHO roadmap or the AAS survey findings. These written comments were imported into NVivo qualitative data analysis package and we undertook a pragmatic thematic content analysis. Analysing the data from the workshops allowed a further open consideration of current research priorities as this step expanded beyond the limitation that the survey had of asking questions within the framework of the WHO roadmap. Following the methodology established after the first workshop (7) we compiled a dataset by transcribing the spoken and written comments from each workshop. A coding framework was generated through an inductive and then deductive approach, following the same categories used in the survey.

#### Patient and public involvement

The participants in this study were The Global Health Research and Health Care community and the very aim was to give them a voice in the requirement to assess whether the right research questions are being tackled in COVID-19. We made ongoing open calls through social media for contributions to surveys and the workshops were open access on The Global Health Network and also on Facebook. The research question was set to address prior lack of engagement with the wider, global community and the design was based on ongoing engagement with this community and our understanding of how to most effectively engageand gain their involvement. The study was entirely open throughout all the steps and the time taken to complete the survey and taking part in the workshops was made clear to participants.

## Results

In total, 1,528 individuals completed the online survey and 2,559 attended the workshops, from across 137 countries, ensuring representation from all of the WHO regions (African region = 612 (40%); Americas region = 279 (18%); Eastern Mediterranean region = 32 (2%); European region = 460 (30%); South East Asia region = 87 (6%); Western Pacific region = 58 (4%)). Participants were most commonly employed in academia (47%), hospitals (14%) research organisations (11%) and non-government organisations (10%).

### Current Global Ranking of the WHO Roadmap Priorities

The survey results (Table 1) shows how priorities were ranked across the immediate and longer term within the WHO categories. We present these globally, along with a sub-group analysis of less resourced countries to understand whether there are differences in priorities for less resourced countries.

**Table 1:**
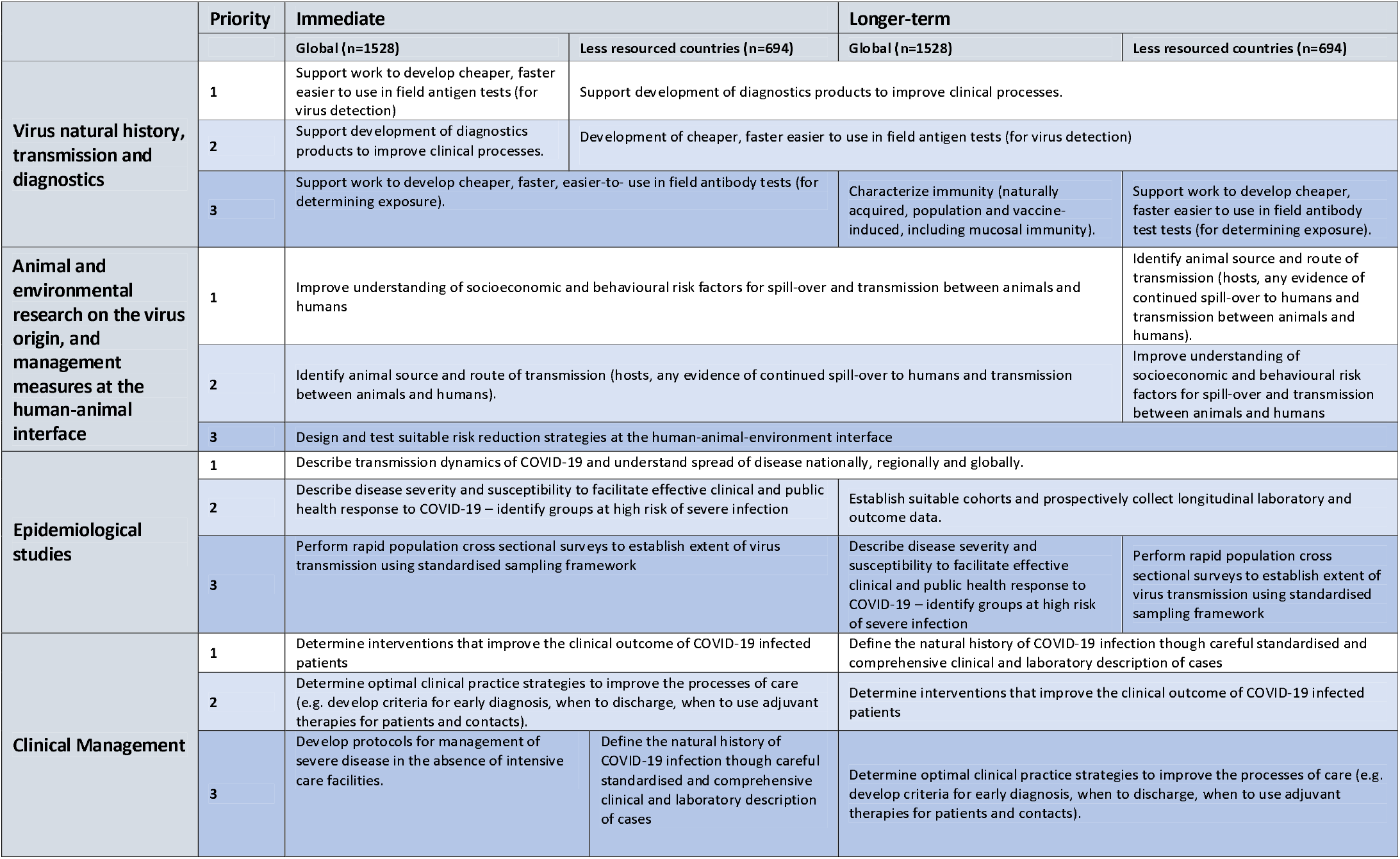
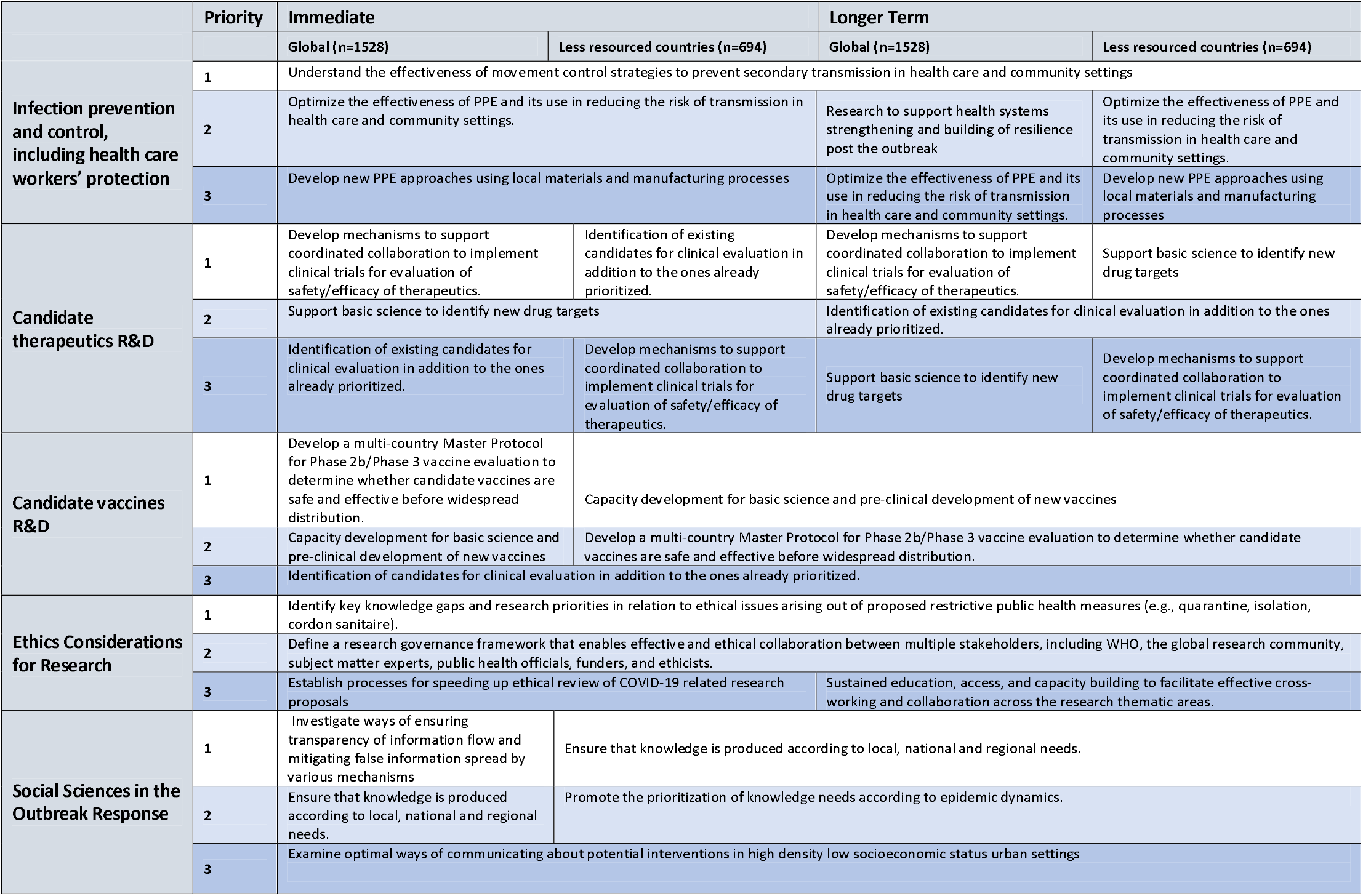
Survey Results: Top three research priorities from the WHO Roadmap categories showing less resources countries as a sub-set of the global responses.

The ranking of these priorities broadly indicates what researchers feel to be the most important research areas from the WHO roadmap at this point within this pandemic. The qualitative data from the survey and the workshops then provides further insight to guide where emphasis should be placed and where completely new priorities are relevant, particularly in low-resourced nations.

The qualitative data analysis from the survey, workshops and working groups supported the existing WHO Roadmap and highlights where greater research emphasis is needed at this later point in the pandemic. However, most importantly new broader priorities have also come through from this study (Table 2).

**Table 2:**
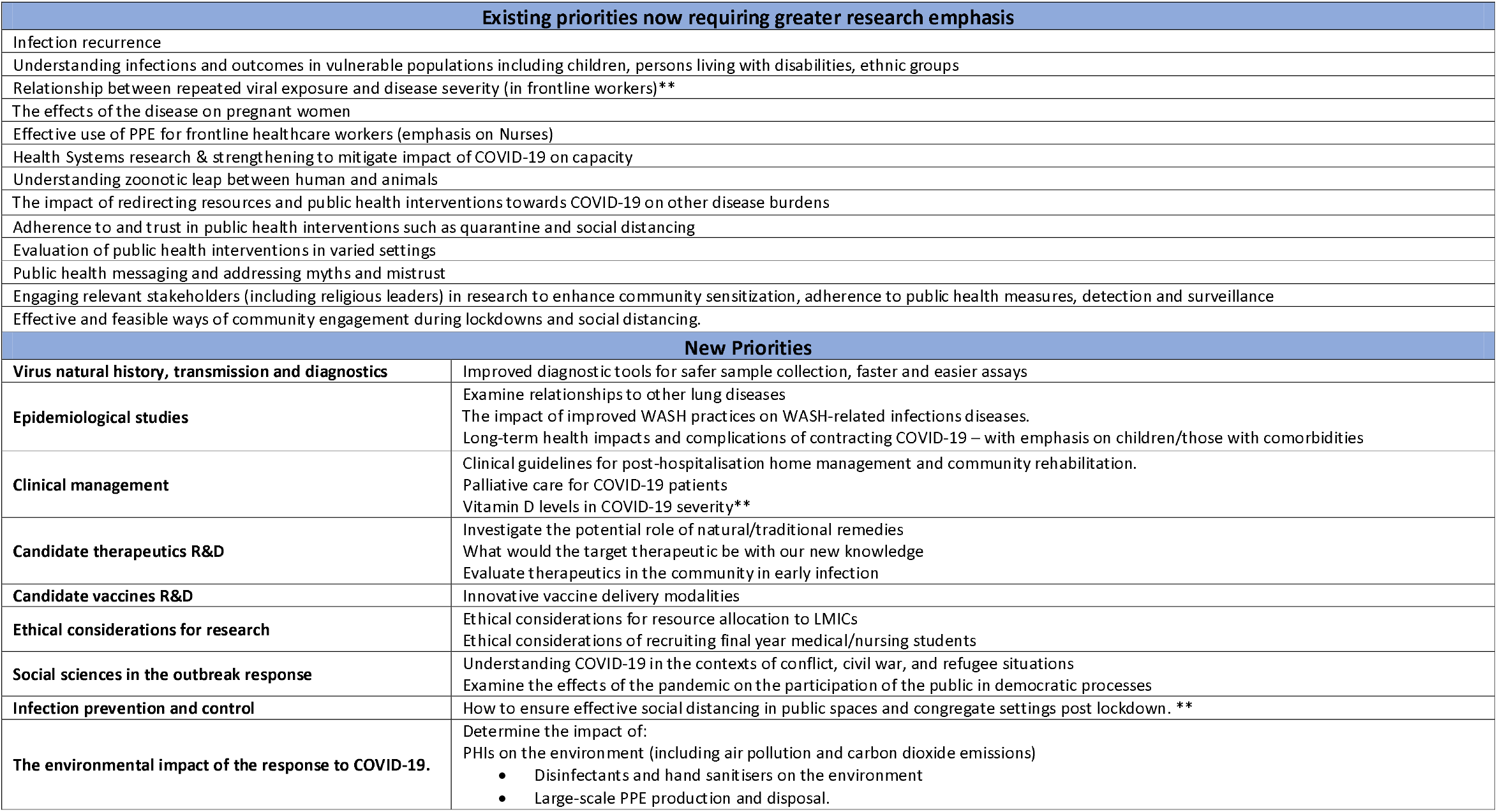
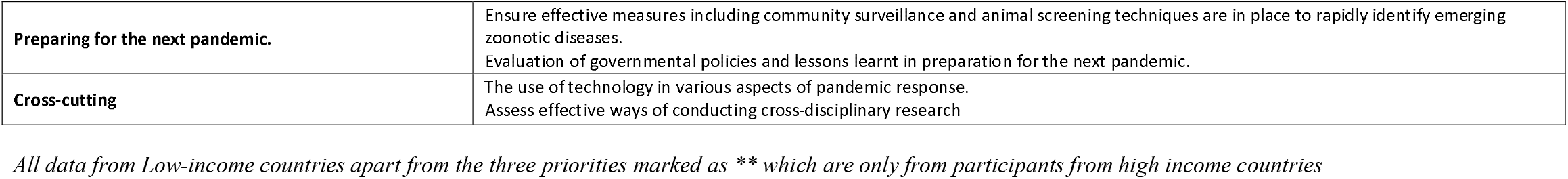
Existing priorities now requiring greater research emphasis and new priorities not in the WHO Roadmap or AAS list (all data from participants working in less resourced countries apart from those priorities asterixed which originated from participants working in higher income countries)

## Discussion

These data suggest that that original WHO COVID-19 Research Roadmap remains broadly globally applicable. Here we also show which research questions require the most emphasis and also that potential new priorities have emerged that were not within the initial roadmap.

Some new suggested priorities reflect the progress of the pandemic and acquisition of knowledge as to where the gaps lie; notably research in children, pregnancy, long-term health impacts of the disease and that there is a strong call for research that assesses the effectiveness of public health measurers put into place across the globe to reduce transmission of this virus. These were alongside a demand for greater social science research to determine public perception, and better ways to change behaviours and build trust (including a need for social sciences to cross-cut the other more biomedical priorities). We also identified a range of new priorities relating to addressing COVID-19 in lower resource settings, where multiple pressures including ongoing endemic infectious diseases and other co-morbidities are competing within the health and policy systems for limited resources. These pressures have led to emphasis on cheaper and field applicable tools and research and health capacity strengthening.

The need for further studies to evaluate public health measures and studies on other potential interventons as they arise were ranked highly by the survey respondents and workshop participants. These studies must be undertaken as quickly as possible, in highly varied social contexts, if we are to gain evidence now on just how effective measures such as lockdown, handwashing and social distancing are on reducing transmission and to understand the relative risks and benefits. The need for social science research and mixed methods came through very strongly, with an emphasis on determining how to gain trust and successfully deliver public health messages. This needs evidence-based community engagement strategies; tested and evaluated everywhere.

Limitations of our approach include the fact that we built the questions to align with the original WHO broad priority headings, this would have inherently focussed the survey respondents around the largely biomedical focus of these priorities and this meant that some headings (for example the animal human interface) had relatively few suggested priorities while others (for example social sciences in the outbreak response) had much larger numbers. We also retained the original order of priorities from the WHO Research Roadmap and the AAS survey and this may have influenced the ranking given by respondents. The workshops however were open and purposefully invited researchers to make whatever comments they wanted in regard to where current research priorities lie, beyond the scope of the WHO Research Roadmap. Therefore, taken together we suggest that these data support the importance of the WHO Research Roadmap approach and highlight where funders and researcher should be placing emphasis as well as identifying potential new areas that should be tackled within this pandemic.

Consideration of both immediate and long-term priorities is important to address this specific pandemic and to better prepare for the future. There are studies that need ongoing transmission, at a high enough rate to answer the question they set. These might be essential for this pandemic, for example clinical trials to determine the efficacy of drugs or vaccines, or address questions to guide future outbreaks, such as evaluating the effectiveness of public health interventions. Other studies do not need circulating virus, and could still guide the effort to address COVID-19 or might help for future pandemics. Fig 1 shows these four situations and gives examples.

Consideration of these findings in the context of where we are now with the global shifting and evolution of the pandemic requires both research teams and funders to ensure research across all these key areas within this finite window. This complements ongoing work by the UK Collaborative on Development Research and Global Research Collaboration for Infectious Disease Preparedness to map research funding against the WHO roadmap priorities to enable funders and researchers identify gaps and opportunities, and inform future research investments or coordination needs.(8)

Finally, we want to highlight both the importance of fully involving the global research community in priority setting, as well as the ongoing need to review priorities where knowledge and practice is advancing rapidly. We recognise that these efforts need to be complemented by further research priority scoping work, beyond the global health focus to further strengthen cross-disciplinary efforts. Here we have shown that the global health research community support the recommendations of the WHO Research Roadmap, but that important new priorities have emerged both due to the transition through the pandemic and consideration of differing global epidemiological, health system, policy, and research contexts.

## Data Availability

All data will be fully available without restriction. URLs/accession numbers/DOIs will be available only after acceptance
of the manuscript for publication so that we can ensure their inclusion before
publication.

## Acknowledgments

We acknowledge and thank all survey and workshop participants for their contributions.

## Footnotes

### Contributors

KM, CA, WM, JM and MA developed the original survey with input from MT, all the authors then contributed to further developing and delivering this global version with oversight from TL. AN and TL guided this analysis along with AD, NC, EA with support from ZA and JP. The workshops were delivered by TL and NF, with support from ZA and JP. TL led the drafting with AN and MT, PP and KM were closely involved throughout and contributed to the draft and review. The other authors contributed significantly and equally in conducting the study and analysing the data. The corresponding author attests that all listed authors meet authorship criteria and that no others meeting the criteria have been omitted. TL is responsible for the overall content as guarantor.

### Funding

The Global Health Network is supported by a grant from the Bill and Melinda Gates Foundation (https://www.gatesfoundation.org/ Grant number: OPP1169808). The COVID-19 Knowledge hub is supported by a grant from UK Research and Innovation (https://www.ukri.org/ Grant number: MC_PC_19073). No other specific funding supported this work. The funders played no role in study design; in the collection, analysis, and interpretation of data; in the writing of the report; or in the decision to submit the paper for publication. All researcher are independent from funders and all authors, external and internal, had full access to all of the data in the study and can take responsibility for the integrity of the data and the accuracy of the data analysis

### Competing interests

All authors have completed the ICMJE uniform disclosure form at www.icmje.org/coi_disclosure.pdf and declare: no support from any organisation for the submitted work; no financial relationships with any organisations that might have an interest in the submitted work in the previous three years; no other relationships or activities that could appear to have influenced the submitted work.

### Ethical approval

This research was limited to seeking the views of healthcare professionals and research staff; patients and the wider community where not involved. Therefore, this research would be considered ‘minimal risk’ and does not come under the definition of research involving human subjects. However, this work does still fall with our research methodology and remit for the protocol that is approved by the University of Oxford Research Ethics Committee (OxTREC) protocol number OxTREC 541-18.

### Data sharing

All the data from this study will be openly available on the global health network.

### Transparency

The lead author (the manuscript’s guarantor) affirms that the manuscript is an honest, accurate, and transparent account of the study being reported; that no important aspects of the study have been omitted; and that any discrepancies from the study as originally planned (and, if relevant, registered) have been explained.

### Dissemination

The results from the survey were shared with the community through the ‘research prioritise’ workshop and the reports from each workshop are being shared on the platform. The wider, cumulative report is being shared online and the release of that will also be widely disseminated. One of our core aims with this research is to make these findings as widely known as possible so that the prioritise that this work highlights translates to studies undertaken by this same community.

### Copyright/license for publication

I, the Submitting Author has the right to grant and does grant on behalf of all authors of the Work (as defined in the below author licence), an exclusive licence and/or a non-exclusive licence for contributions from authors who are: i) UK Crown employees; ii) where BMJ has agreed a CC-BY licence shall apply, and/or iii) in accordance with the terms applicable for US Federal Government officers or employees acting as part of their official duties; on a worldwide, perpetual, irrevocable, royalty-free basis to BMJ Publishing Group Ltd (“BMJ”) its licensees and where the relevant Journal is co-owned by BMJ to the co-owners of the Journal, to publish the Work in BMJ Global Health and any other BMJ products and to exploit all rights, as set out in our licence.

## Abbreviations

WHO: World Health Organisation
AAS: African Academy of Science
TGHN: The Global Health Network

---

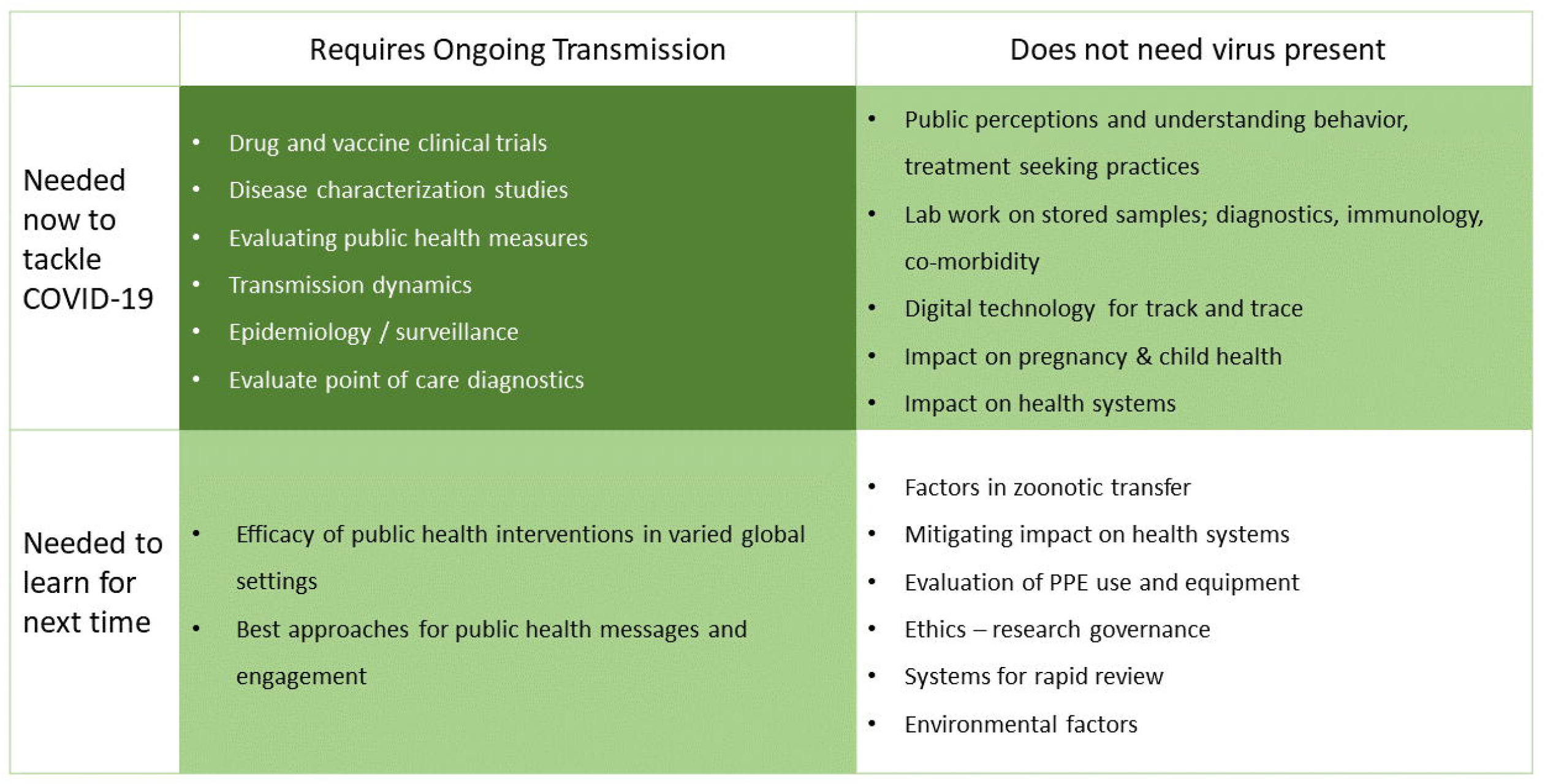

## Notes

### Competing Interest Statement

The authors have declared no competing interest.

## References

1. World Health Organization. WHO Director-General’s statement on IHR Emergency Committee on Novel Coronavirus (2019-nCoV) [cited 2020 June 6]. [Internet]. Available from: https://www.who.int/dg/speeches/detail/who-director-general-s-statement-on-ihr-emergency-committee-on-novel-coronavirus-(2019-ncov).

2. World Health Organization. WHO Director-General’s opening remarks at the media briefing on COVID-19 - 11 March 2020 [cited 2020 June 11]. [Internet]. Available from: https://www.who.int/dg/speeches/detail/who-director-general-s-opening-remarks-at-the-media-briefing-on-covid-19---11-march-2020.

3. World Health Organization. A COORDINATED GLOBAL RESEARCH ROADMAP: 2019 NOVEL CORONAVIRUS Geneva: World Health Organization; [cited 2020 June 11]. [Internet]]. Available from: https://www.who.int/blueprint/priority-diseases/key-action/Coronavirus_Roadmap_V9.pdf?ua=1.

4. Lang T. Ebola: Embed research in outbreak response. Nature. 2015;524(7563):29–31.

5. Piot P, Soka MJ, Spencer J. Emergent threats: lessons learnt from Ebola. Int Health. 2019;11(5):334–7.

6. The African Academy of Sciences. Research and Development goals for COVID-19 in Africa - The African Academy of Sciences Priority Setting Exercise [cited 2020 June 11]. Available from: https://www.aasciences.africa/sites/default/files/2020-04/Research%20and%20Development%20Goals%20for%20COVID-19%20in%20Africa.pdf.

7. Tibenderana J, Alia J, Hamade P, Walker R, Feune De Colombi N, Al-Rawni Z, et al. Malaria and COVID-19: A rapid determination of unknowns and call for research. Paper finalised MedRxiv Pre-print server on 10^th^ July 2020.

8. The UK Collaborative on Development Research. COVID-19 Research Project Tracker by UKCDR & GloPID-R [cited 2020 June 11]. [Internet]. Available from: https://www.ukcdr.org.uk/funding-landscape/covid-19-research-project-tracker/.

